# Large Language Model–Enhanced Electronic Frailty Assessment and Prognostic Performance in Hospitalized Adults: A Retrospective Cohort Study

**DOI:** 10.64898/2026.11.04.26367272

**Authors:** Anuj K. Dalal, John Lundstedt, Natalie C. Ernecoff, Savanna Plombon, Shoshana Streiter, Marie Leeson, Joseph M. Plasek, Stephanie Mueller, Julie Fiskio, Li Zhou, Stuart Lipsitz, Ariela Orkaby, Julia V. Loewenthal

## Abstract

**Background:** Electronic frailty indices (eFIs) derived from structured electronic health record (EHR) data may incompletely capture functional deficits. Large language models (LLMs) can extract frailty-related information from clinical notes, but whether improved frailty ascertainment improves prognostic performance is uncertain.

**Objective:** To compare frailty ascertainment and prognostic performance between standard and LLM-enhanced eFIs among hospitalized adults.

**Design:** Retrospective cohort study

**Participants:** Adults aged ≥50 years enrolled in two studies at an academic medical center and affiliated community hospital between February 2022 and April 2024.

**Main Measures:** A validated 35-deficit diagnosis-based eFI and an LLM-enhanced eFI incorporating deficits extracted from physical therapy (PT) documentation using GPT-4o. Frailty was defined using an index threshold of ≥0.20. The primary outcome was a composite of discharge to a facility, readmission, or death within 90 days. Multivariable logistic regression models assessed associations with the outcome, and discrimination was compared using area under the receiver operating characteristic curve (AUROC). Secondary analyses examined threshold-based classification and PT-documentation availability; prespecified sensitivity analyses evaluated population and outcome restrictions.

**Results:** Among 616 hospitalizations, mean (SD) age was 70.9 (10.2) years, 329 (53.4%) had PT documentation, and 320 (51.9%) experienced the composite outcome. Standard and LLM-enhanced eFIs classified 216 (35.1%) and 362 (58.8%) hospitalizations as frail, respectively. Both were independently associated with the outcome (standard eFI: adjusted OR, 1.98 [95% CI, 1.39–2.83]; LLM-enhanced eFI: adjusted OR, 2.46 [95% CI, 1.75–3.47]). Discrimination did not differ significantly (AUROC, 0.636 vs 0.660; difference, 0.024 [95% CI, −0.006 to 0.053]; *p*=0.12). LLM enhancement increased sensitivity (42.5% to 69.4%) but decreased specificity (73.0% to 52.7%).

**Conclusions:** LLM enhancement substantially increased frailty ascertainment and sensitivity for adverse outcomes but did not significantly improve prognostic discrimination. Thus, LLM-enhanced frailty assessment may depend on whether the intended use prioritizes broad identification of functional vulnerability or prediction of adverse post-hospital outcomes.

## INTRODUCTION

Frailty is a multidimensional syndrome of decreased physiologic reserve resulting from accumulated deficits across health domains and is associated with adverse outcomes—such as readmission, ongoing facility care, and mortality—among hospitalized older adults.^1–4^ Because frailty captures vulnerability beyond chronologic age,^5,6^ identifying frailty during hospitalization may inform risk stratification and targeted care of vulnerable adults. Frailty screening is increasingly relevant to inpatient care, reinforced by national initiatives such as the Centers for Medicare & Medicaid Services Age-Friendly Hospital Measure.^3,7–9^

Comprehensive geriatric assessment can characterize frailty but is resource intensive and difficult to implement universally.^8,10–15^ Electronic frailty indices (eFIs), which quantify accumulated deficits using passively collected electronic health record (EHR) data, offer a validated alternative.^16–19^ However, eFIs based primarily on structured diagnosis codes may incompletely capture functional deficits, including mobility impairment, gait abnormalities, and falls, that are often documented in narrative clinical notes, such as physical therapy (PT) documentation.^8,20,21^

Large language models (LLMs) provide a scalable means of extracting frailty-related information from unstructured documentation.^8,22^ In prior work, we developed and validated an LLM-enhanced eFI incorporating deficits extracted from PT notes.^23^ Compared with the standard eFI, the LLM-enhanced eFI demonstrated greater discrimination and sensitivity for frailty defined by a geriatrician-performed comprehensive geriatric assessment.^23^

Whether improved frailty ascertainment translates into improved prognostic stratification remains uncertain.^9^ Frailty measurement and prediction of post-hospital outcomes are distinct objectives, and additional functional information may identify more patients as frail without proportionately improving prediction of outcomes influenced by acute illness, social factors, and transitions of care. We therefore compared standard and LLM- enhanced eFIs among at-risk hospitalized middle-aged and older adults, evaluating their associations with and discrimination of adverse post-hospitalization outcomes.^8,24,25^ Secondarily, we examined changes in frailty classification and threshold-based prognostic performance. We hypothesized that LLM augmentation would identify more patients as frail and that frailty identified by either approach would be associated with adverse outcomes.

## METHODS

### Study Design, Setting, and Population

We conducted a secondary, retrospective analysis of two cohorts of at-risk adults hospitalized between February 2022 and April 2024 at Brigham and Women’s Hospital and Brigham and Women’s Faulkner Hospital within Mass General Brigham (MGB), Boston, Massachusetts. Study 1 enrolled general medicine patients with two or more chronic conditions discharged home,^27^ and Study 2 enrolled patients undergoing interhospital transfer.^28^ We restricted the analytic cohort to adults aged ≥50 years because frailty and functional vulnerability are present in middle age and potentially associated with adverse outcomes following acute hospitalization.^5,6,29,30^ Detailed eligibility criteria were previously reported. ^27,28^

We followed TRIPOD+AI and applicable TRIPOD-LLM reporting guidance. ^26^ Because we evaluated prespecified frailty indices rather than developed a prediction model, model-development items were not applicable (**Supplement**). Consent procedures for each source study were approved by the MGB Institutional Review Board.

### Data Sources and Covariates

Demographic, clinical, and outcome data were obtained from the electronic health record (EHR; Epic Systems, Inc.) and institutional enterprise data warehouse (EDW).

Prespecified covariates included age, sex, race and ethnicity, marital status, primary language, education, insurance type, employment status, and source-study cohort. These covariates were selected *a priori* based on their established associations with post-discharge outcomes.^31–34^ We intentionally did not include individual comorbidities or a separate comorbidity index because morbidity-related deficits are constituent components of the frailty measures (below) under evaluation. Structured data included International Classification of Diseases, Tenth Revision (ICD-10) diagnosis codes from the 24 months preceding index hospitalization; unstructured data consisted of PT documentation from index hospitalization.

### Frailty Measures

#### Standard Electronic Frailty Index

Frailty was assessed using a previously validated MGB eFI comprising 35 deficits across morbidity, functional, sensory, and cognitive domains.^35^ Deficits were identified from ICD-10 codes during the preceding 24 months, and the eFI was calculated as the number of deficits present divided by 35. Frailty was defined as eFI ≥0.20.

#### LLM-Enhanced Electronic Frailty Index

The previously developed and validated LLM-enhanced eFI supplements the standard eFI with frailty-related deficits extracted from PT documentation and demonstrated greater discrimination and correlation with geriatrician-assessed frailty than the standard eFI.^23^ For this study, GPT-4o processed PT documentation in a HIPAA-compliant Microsoft Azure environment using standardized prompts to identify the same 35 deficits. The prior validation study used the element-wise maximum of structured and LLM-derived deficits to construct the enhanced eFI, consistent with this approach.^23^

For each deficit, the enhanced eFI used the maximum value identified from structured ICD-10 data or LLM-derived information, thereby adding deficits documented in PT notes but absent from structured diagnoses. Frailty scores and the ≥0.20 threshold were otherwise calculated identically to the standard eFI. For hospitalizations without PT documentation, the enhanced eFI equaled the standard eFI. Prompt specifications and operational definitions are provided in the **Supplement** and were described previously.^23^

### Outcomes

The primary outcome was a composite of hospital readmission, discharge to a post- acute or long-term care facility following the index or a subsequent hospitalization, or death, occurring at discharge or within 90 days. Facility discharge was included as a marker of vulnerability and hospitalization-associated functional decline.^30,36–38^ Functional vulnerability, in particular, may be associated with adverse outcomes among middle-aged adults with multimorbidity and unrecognized frailty following acute hospitalization.^5,25^ Outcomes were ascertained from EHR data consistently across both source studies and independently of frailty classification.

### Sample

Based on prior validation, we anticipated AUROCs of approximately 0.62 for the standard eFI and 0.70 for the LLM-enhanced eFI. With approximately 600 eligible hospitalizations from the source study cohorts and a balanced outcome distribution, the available sample was expected to detect an AUROC difference of approximately 0.08; smaller differences were examined under varying assumptions regarding correlation between paired predictions.

### Statistical Analysis

Patient characteristics, PT-documentation availability, frailty measures, and outcomes were summarized using mean (SD) for continuous variables and counts (percentages) for categorical variables. Differences by composite-outcome status were assessed using Pearson chi-square or Fisher exact tests, as appropriate.

Primary analyses used separate multivariable logistic regression models containing either the standard or LLM-enhanced eFI (frailty threshold ≥0.20), adjusted for age, sex, race and ethnicity, marital status, primary language, education, insurance, employment status, and source study. We reported adjusted odds ratios (ORs) with 95% CIs. Missing categorical values were retained as a separate category without imputation. Model discrimination was assessed using AUROC with 95% CIs and compared using DeLong’s test.

Secondary analyses compared sensitivity, specificity, positive and negative predictive values, positive and negative likelihood ratios, accuracy, balanced accuracy, and F1-score at eFI thresholds of ≥0.20 and ≥0.30. These measures assessed classification of the adverse outcome rather than diagnostic accuracy for frailty, which was previously evaluated against a geriatrician-performed reference standard.^23^ Because PT documentation could reflect functional assessment and discharge planning, we repeated both models with adjustment for PT-documentation availability and evaluated effect modification using an interaction between LLM-enhanced frailty and PT-documentation availability.

Sensitivity analyses repeated the primary models among hospitalizations with PT documentation, adults aged ≥65 years, patients discharged home after the index hospitalization, and separately within each source-study cohort. We also repeated all analyses using an alternative composite outcome of hospital readmission or death within 90- days, excluding facility discharge from the outcome definition.

### Exploratory Manual Chart Review

In a random 10% subset of hospitalizations, comprehensive EHR review assessed incremental deficit ascertainment associated with LLM augmentation of the standard eFI. Manual review incorporated the structured diagnoses used to derive the standard eFI together with PT, occupational therapy, case management, social work, and physician documentation. This analysis was intended to characterize additional deficits identified through LLM augmentation and their confirmation on comprehensive EHR review, rather than to estimate accuracy against an independent reference standard. Two trained reviewers independently evaluated a subset using standardized abstraction procedures, with clinician adjudication of discrepancies; Cohen’s κ >0.80 was established before subsequent single-reviewer abstraction. Detailed deficit definitions and chart-review procedures are provided in the **Supplement**.

## RESULTS

### Cohort Characteristics, Frailty Classification, and Outcomes

Of 795 hospitalizations identified from the two source cohorts (293 from Study 1; 502 from Study 2), 179 were excluded (101 from Study 1, 78 from Study 2) because patients were aged <50 years. A total of 616 hospitalizations (601 unique patients) were available for analysis: 192 (31.2%) were from Study 1 and 424 (68.8%) from Study 2. Mean (SD) age was 70.9 (10.2) years, 272 (44.2%) patients were female, and 329 (53.4%) hospitalizations had PT documentation (**Table 2**).

**Table 1.** Summary of Regression Models and Analytic Objectives.

| Model | Key Exposures | Population | Purpose |
|---|---|---|---|
| *Primary Analysis* |  |  |  |
| A | Standard eFI (≥0.2) | Full cohort | Evaluate association with composite outcome. |
| B | LLM-enhanced eFI (≥0.2) | Full cohort | Evaluate association with composite outcome and compare discrimination with Model A |
| *Secondary Analysis* |  |  |  |
| C | Standard eFI (≥0.2), PT note availability | Full cohort | Evaluate association after accounting for PT-documentation availability |
| D | LLM-enhanced eFI (≥0.2), PT note availability | Full cohort | Evaluate association after accounting for PT-documentation availability |
| E | LLM-enhanced eFI (≥0.2), LLM-enhanced eFI × PT availability interaction | Full cohort | Evaluate whether PT availability modifies the association between LLM-enhanced frailty ascertainment and the composite outcome |
| *Sensitivity Analyses* |  |  |  |
| F | Models A and B | PT note subset | Repeat primary analyses in cases with available PT documentation |
| G | Models A and B | Adults ≥65 years | Examine robustness in older adults |
| H | Models A and B | Home disposition | Exclude index hospitalization facility discharges |
| J | Models A and B | Alternative outcome definition | Repeat all analyses with outcome defined as 90-day readmission or mortality. |
| I | Models A and B | Source study | Examine consistency across study populations |

All models included adjustment for the following covariates: age, sex, ethnicity, language, race, marital status, employment, insurance, and source study.

**Table 2.** Cohort Characteristics by Composite Outcome Status.

| **Characteristic** | **N** | **Outcome, n (%)** | **No outcome, n (%)** | ***p-value*** |
|---|---|---|---|---|
| **Hospitalizations** | 616 | 320 (51.9) | 296 (48.1) | - |
| **Unique patients** | 601 | 308 (51.2) | 293 (48.8) | - |
| **Source cohort** |  |  |  |  |
| Study 1 (ePROs) | 192 | 93 (48.4) | 99 (51.6) | 0.241 |
| Study 2 (IHT) | 424 | 227 (53.5) | 197 (46.5) |  |
| **Age** |  |  |  |  |
| <65 years | 179 | 88 (49.2) | 91 (50.8) | 0.376 |
| ≥65 years | 437 | 232 (53.1) | 205 (46.9) |  |
| **Gender** |  |  |  |  |
| Female | 272 | 146 (53.7) | 126 (46.3) | 0.45 |
| Male | 344 | 174 (50.6) | 170 (49.4) |  |
| **Race and ethnicity** |  |  |  |  |
| White, non-Hispanic | 455 | 237 (52.1) | 218 (47.9) | 0.794 |
| Black, non-Hispanic | 89 | 49 (55.1) | 40 (44.9) |  |
| Hispanic | 33 | 16 (48.5) | 17 (51.5) |  |
| Other/missing | 39 | 18 (46.2) | 21 (53.8) |  |
| **Marital status** |  |  |  |  |
| Partnered | 298 | 134 (45.0) | 164 (55.0) | **<0.01** |
| Not partnered | 313 | 185 (59.1) | 128 (40.9) |  |
| Missing | 5 | 1 (20.0) | 4 (80.0) |  |
| **Primary language** |  |  |  |  |
| English | 577 | 305 (52.9) | 272 (47.1) | 0.08 |
| Non-English | 39 | 15 (38.5) | 24 (61.5) |  |
| **Education** |  |  |  |  |
| Less than high school | 44 | 28 (63.6) | 16 (36.4) | 0.20 |
| High school/GED | 250 | 133 (53.2) | 117 (46.8) |  |
| Some college | 15 | 10 (66.7) | 5 (33.3) |  |
| College/higher education | 239 | 119 (49.8) | 120 (50.2) |  |
| Missing | 68 | 30 (44.1) | 38 (55.9) |  |
| **Insurance** |  |  |  |  |
| Public | 320 | 179 (55.9) | 141 (44.1) | 0.10 |
| Private | 281 | 135 (48.0) | 146 (52.0) |  |
| Other | 15 | 6 (40.0) | 9 (60.0) |  |
| **Employment** |  |  |  |  |
| Employed | 134 | 61 (45.5) | 73 (54.5) | 0.35 |
| Unemployed | 82 | 43 (52.4) | 39 (47.6) |  |
| Retired | 314 | 172 (54.8) | 142 (45.2) |  |
| Other/missing | 86 | 44 (51.2) | 42 (48.8) |  |
| **PT documentation** |  |  |  |  |
| Available | 329 | 202 (61.4) | 127 (38.6) | **<0.01** |
| Not available | 287 | 118 (41.1) | 169 (58.9) |  |
| **Standard eFI** |  |  |  |  |
| ≥0.20 | 216 | 136 (63.0) | 80 (37.0) | **<0.01** |
| <0.20 | 400 | 184 (46.0) | 216 (54.0) |  |
| **LLM-enhanced eFI** |  |  |  |  |
| ≥0.20 | 362 | 222 (61.3) | 140 (38.7) | **<0.01** |
| <0.20 | 254 | 98 (38.6) | 156 (61.4) |  |

Data are presented as counts (N) and n (row %) unless otherwise indicated. Row percentages indicate the proportion of hospitalizations within each characteristic category with or without the composite outcome. P-values were calculated using Pearson's chi-square test, with missing values included as separate categories where indicated. Exact tests were used when expected cell counts were small. Analyses were conducted at the hospitalization level (N = 616), representing 601 unique patients.

The composite outcome occurred following 320 hospitalizations (51.9%), including 226 readmissions, 145 facility discharges, and 53 deaths (outcome sub-components were not mutually exclusive). Of the 145 facility discharges, 120 occurred after the index hospitalization and 25 after subsequent hospitalization.

At the prespecified ≥0.20 threshold, the standard eFI classified 216 hospitalizations (35.1%) as frail, whereas the LLM-enhanced eFI classified 362 (58.8%) as frail. Among hospitalizations with the composite outcome (n=320), 42.5% and 69.4% were classified as frail, respectively; corresponding percentages among those without the outcome (n=296) were 27.0% and 47.3%, respectively.

### Primary Analysis

Frailty was independently associated with the composite outcome using both the standard eFI (adjusted OR, 1.98; 95% CI, 1.39–2.83) and LLM-enhanced eFI (adjusted OR, 2.46; 95% CI, 1.75–3.47). Model discrimination was similar (AUROC, 0.636 vs 0.660; difference, 0.024 [95% CI, −0.006 to 0.053]; *p*=0.12) (**Figures 1 and 2**)

**Figure 1.**
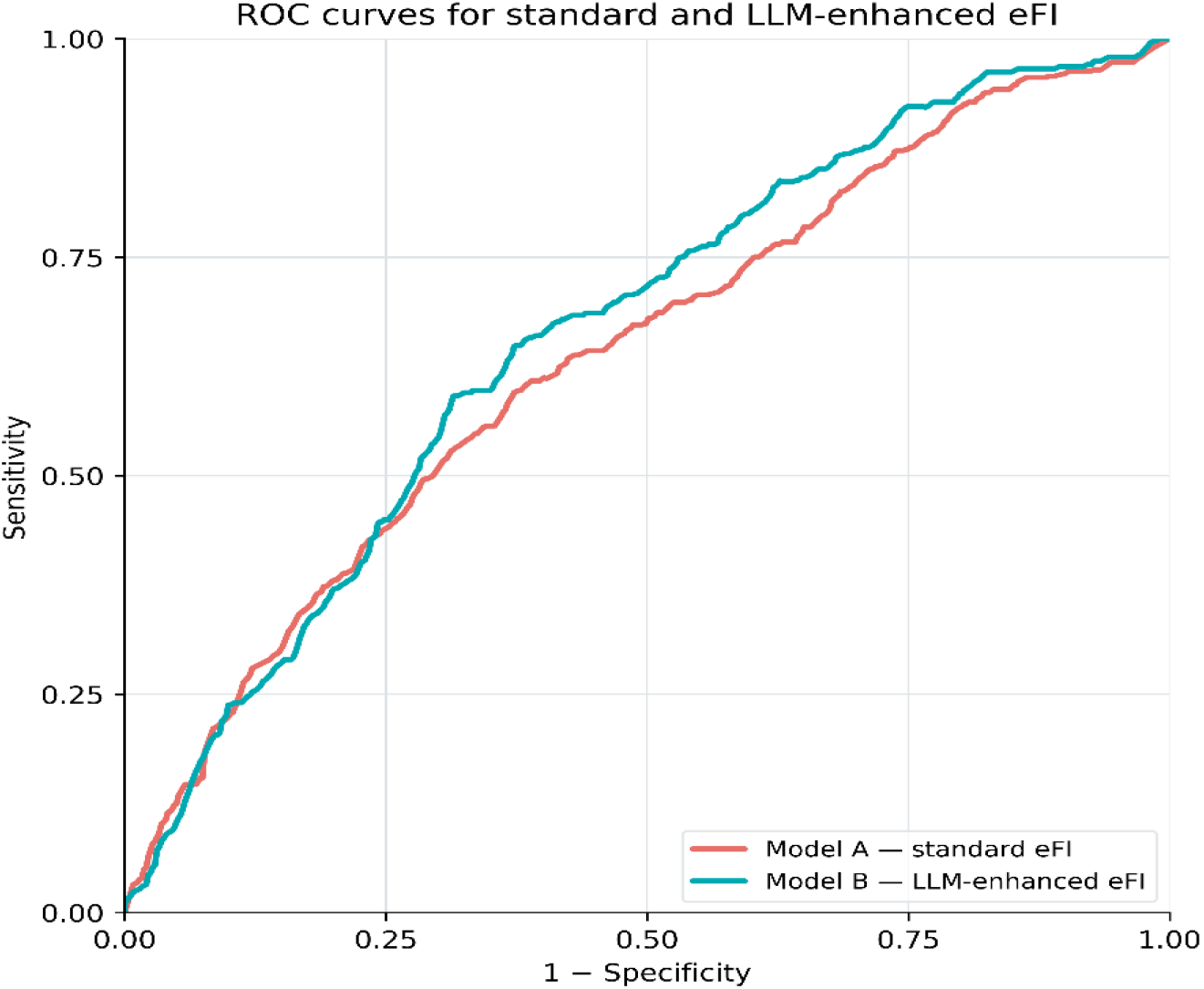
ROC Curves for Standard and LLM-Enhanced Electronic Frailty Indices. Receiver operating characteristic (ROC) curves compare prognostic discrimination of adjusted models incorporating the standard electronic frailty index (eFI) and LLM-enhanced eFI for the composite 90-day outcome. AUROCs were 0.636 and 0.660, for Model A and B, respectively; the difference was 0.024 (95% CI, −0.006 to 0.053; *p*=0.12).

**Figure 2.**
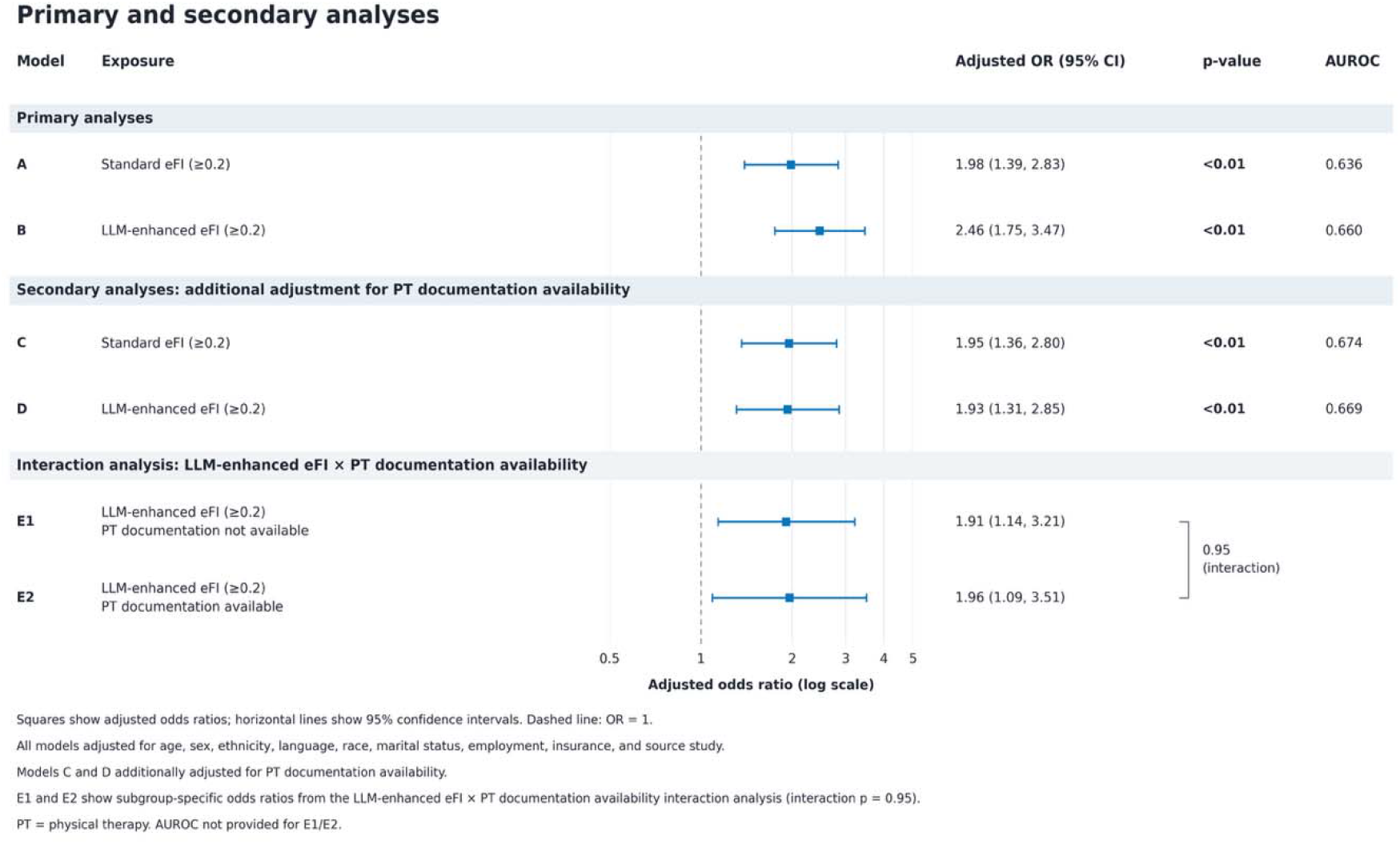
Associations of Standard and LLM-Enhanced Electronic Frailty Indices with the Composite Outcome. Forest plot showing adjusted odds ratios (ORs) and 95% confidence intervals (CIs) from multivariable logistic regression models evaluating associations of the standard electronic frailty index (eFI) and large language model (LLM)-enhanced eFI with the composite outcome. All models were adjusted for age, sex, race and ethnicity, primary language, marital status, education, insurance, employment status, and source-study cohort; models C and D were also adjusted for PT documentation availability.

### Secondary Analyses

The composite outcome occurred following 61.4% of hospitalizations with PT documentation versus 41.1% without PT documentation. After adjustment for PT- documentation availability, frailty remained associated with the composite outcome using the standard eFI (adjusted OR, 1.95; 95% CI, 1.36–2.80) and LLM-enhanced eFI (adjusted OR, 1.93; 95% CI, 1.31–2.85). In the interaction model, LLM-enhanced frailty remained associated with the composite outcome both among hospitalizations without PT documentation (adjusted OR, 1.91; 95% CI, 1.14–3.20) and with PT documentation (adjusted OR, 1.96; 95% CI, 1.09–3.51), with no evidence of effect modification by PT- documentation availability (interaction p-value = 0.95)

At the ≥0.20 threshold, LLM augmentation increased sensitivity for the composite outcome from 42.5% to 69.4% and decreased specificity from 73.0% to 52.7%. NPV, accuracy, and F1-score were higher with the LLM-enhanced eFI, whereas PPVs were similar. At ≥0.30, sensitivity remained higher with the LLM-enhanced eFI (42.5% vs 21.6%) and specificity lower (74.3% vs 87.8%) (**Table 3**).

**Table 3.**
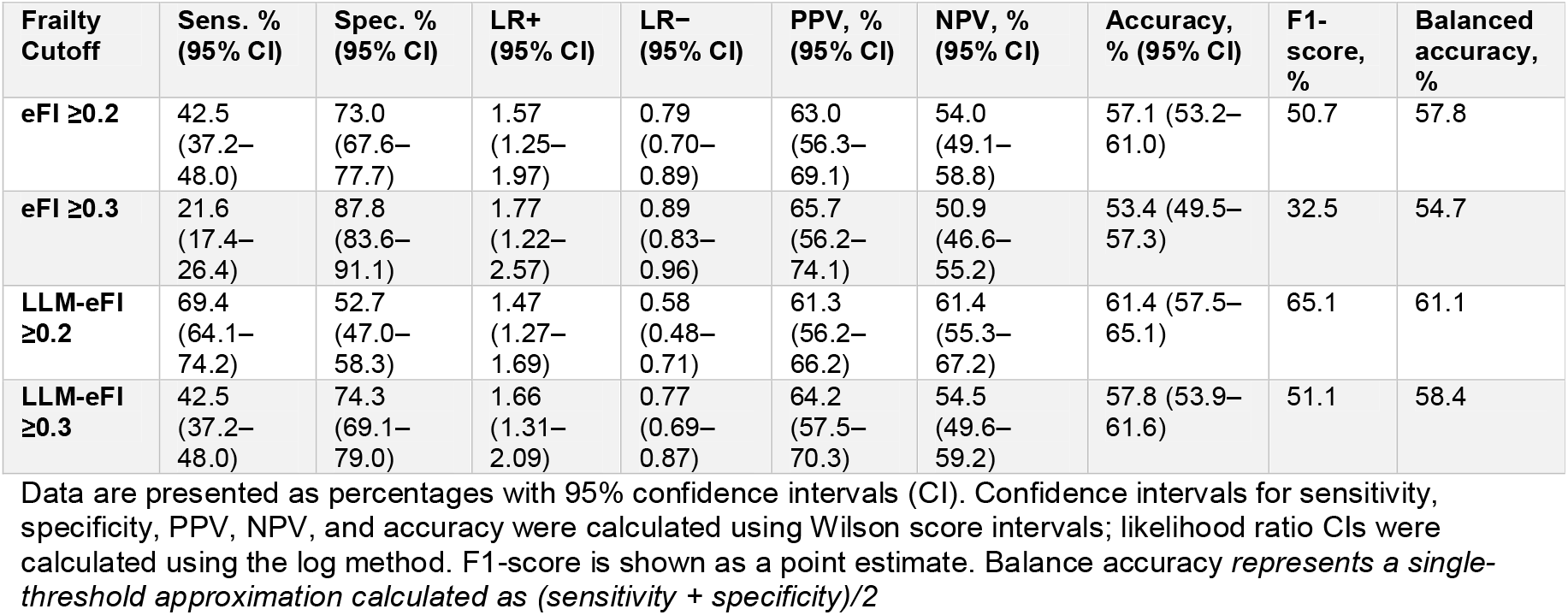
Threshold-based prognostic classification of electronic frailty models across. —f**ull cohort, n=616**

### Sensitivity Analyses

Among the 329 hospitalizations with PT documentation, both measures remained associated with the composite outcome (standard eFI: adjusted OR, 2.25 [95% CI, 1.36– 3.78]; LLM-enhanced eFI: adjusted OR, 2.18 [95% CI, 1.21–3.98]), with AUROCs of 0.656 and 0.631, respectively. At the ≥0.20 threshold, LLM augmentation increased sensitivity from 43.6% to 86.1% but decreased specificity from 71.7% to 24.4% (**Supplemental Table S2**).

Associations remained consistent across additional sensitivity analyses (**Figure 3**). Among adults aged ≥65 years, adjusted ORs were 1.99 and 2.55, with AUROCs of 0.643 and 0.657 for the standard and LLM-enhanced eFIs, respectively. Among patients discharged home, adjusted ORs were 2.33 and 2.28, with an AUROC of 0.658 for both eFIs. When the outcome was alternatively defined as 90-day readmission or mortality, adjusted ORs were 2.52 and 2.49, with AUROCs of 0.646 and 0.649, respectively.

**Figure 3.**
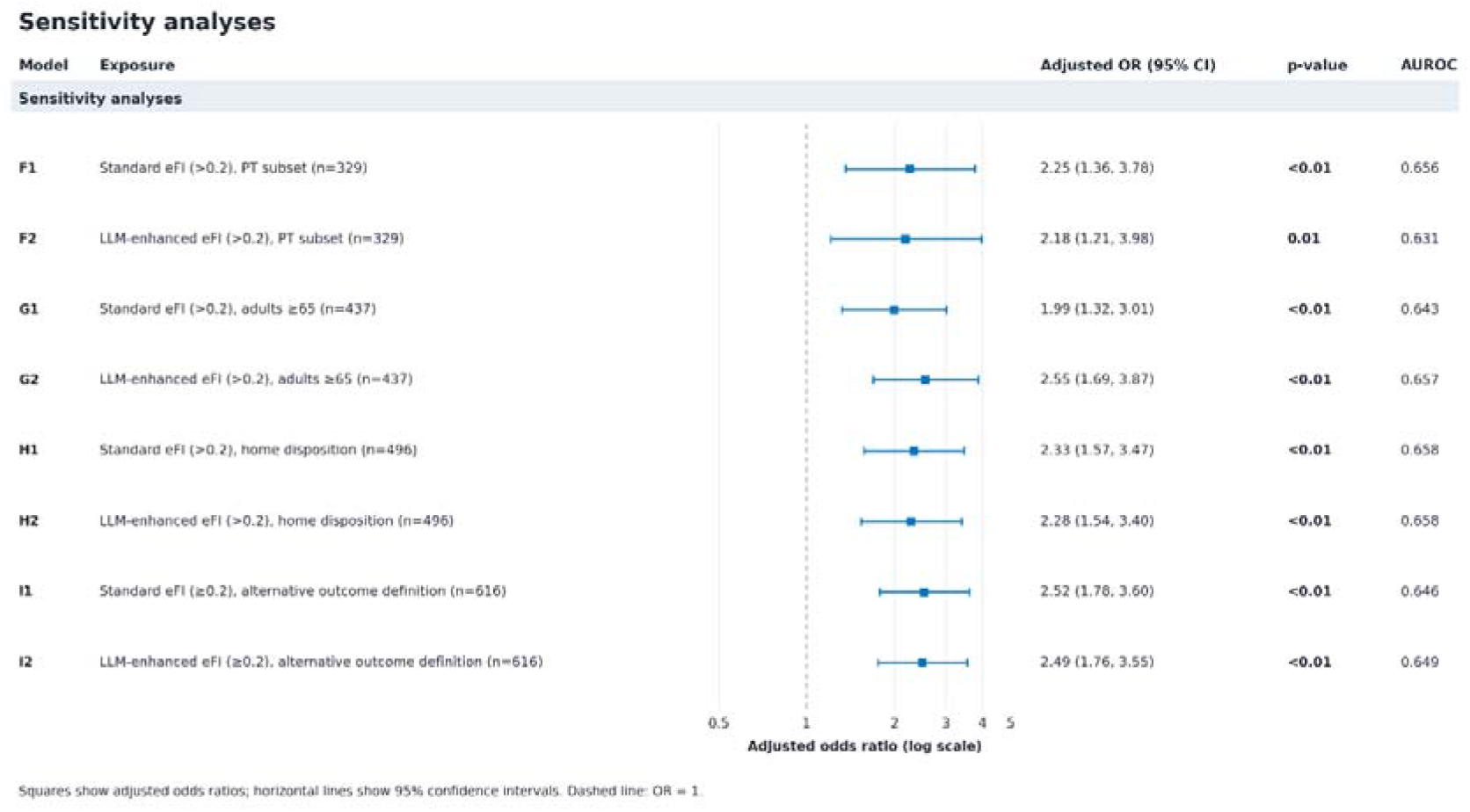
Sensitivity Analyses of Standard and LLM-Enhanced Electronic Frailty Indices. Forest plot showing adjusted odds ratios (ORs) and 95% confidence intervals (CIs) for the association between frailty and the composite adverse outcome in prespecified sensitivity analyses. Separate multivariable logistic regression models evaluated frailty defined by the standard electronic frailty index (eFI) and large language model (LLM)-enhanced eFI at the prespecified threshold of ≥0.20. Sensitivity analyses included restriction to hospitalizations with available PT documentation (n=329), adults aged ≥65 years (n=437), and patients discharged home after the index hospitalization (n=496), as well as an alternative outcome definition comprising 90-day readmission or mortality and excluding facility discharge (n=616). All models were adjusted for age, sex, race and ethnicity, primary language, marital status, education, insurance, employment status, and source-study cohort. Squares indicate adjusted ORs, horizontal lines indicate 95% CIs, and the dashed vertical line indicates an OR of 1. AUROC indicates area under the receiver operating characteristic curve.

Frailty-outcome associations also persisted within each source study, although estimates were less precise. Adjusted ORs for the standard and LLM-enhanced eFIs were 2.44 and 2.23 in Study 1 and 1.96 and 2.66 in Study 2, with corresponding AUROCs of 0.730 and 0.730 in Study 1 and 0.622 and 0.661 in Study 2, respectively (**Supplemental Table S3**).

### Exploratory Manual Chart Review

In the random 10% chart-review sample (n=60), comprehensive manual EHR review identified 830 deficit-level positives. The standard eFI identified 339 (40.8%) of these deficits. Incorporation of LLM-extracted information from PT documentation identified an additional 106 manually confirmed deficits, increasing the number detected to 445 (53.6%) and reducing false negatives from 491 to 385. Of 152 deficit-level positives added through LLM augmentation, 106 (69.7%) were confirmed by comprehensive manual review and 46 (30.3%) were not confirmed. Unconfirmed LLM-derived deficits were concentrated in gait abnormality, cachexia/failure to thrive/debility, falls, fatigue, and use of health services. See **Supplemental Table S4**.

## DISCUSSION

In this retrospective analysis of a cohort of middle-aged and older hospitalized adults, LLM augmentation increased the percentage classified as frail from 35.1% to 58.8%. Frailty defined by either approach was independently associated with the composite adverse outcome, but LLM augmentation did not significantly improve prognostic discrimination (AUROC, 0.636 vs 0.660; *p*=0.12). Instead, augmentation primarily increased sensitivity while decreasing specificity. These findings distinguish two related but different properties of electronic frailty measurement: frailty identification and prediction of adverse outcomes.^39–41^ In our prior study, the LLM-enhanced eFI improved discrimination of geriatrician-assessed frailty (AUROC, 0.701 vs 0.614; *p*<0.001) and correlation with a comprehensive geriatric assessment.^23^ The present findings suggest that more complete frailty ascertainment does not necessarily produce comparable gains in prognostic discrimination. The clinical value of enhanced electronic phenotyping therefore depends on whether its intended purpose is identification, prediction, or informing clinical decisions.^42,43^

Several factors may explain this divergence. Structured eFI data may already capture much of the prognostic information associated with accumulated health deficits, even when functional deficits are incompletely documented. Frailty and multimorbidity frequently coexist but represent distinct constructs associated with adverse outcomes.^44,45^ A meta-analysis found that approximately 72% of frail individuals had multimorbidity, whereas only 16% of individuals with multimorbidity were frail.^44^ Additional deficits identified by LLM augmentation may therefore provide limited independent prognostic information beyond structured diagnoses. Moreover, inpatient PT assessments may reflect acute functional decline as well as baseline frailty, though our operational definitions reflected this distinction. While the source study cohorts represented different inpatient contexts, including interhospital transfer with potentially distinct determinants of adverse outcomes,^46,47^ both eFIs remained associated with post-hospitalization outcomes after adjustment for source study, with consistent findings across multiple sensitivity analyses.

Threshold-based analyses further illustrate the distinction between frailty identification and prognostic performance. At ≥0.20, LLM augmentation increased sensitivity for the composite outcome from 42.5% to 69.4% while decreasing specificity from 73.0% to 52.7%, with a more pronounced tradeoff among hospitalizations with PT documentation.

Exploratory chart review suggested that this broader classification reflected, at least in part, recovery of clinically documented deficits not captured by structured diagnoses. Among deficit classifications added through LLM augmentation (**Supplemental Table S5**), most were confirmed on comprehensive EHR review, although some were not, indicating a tradeoff between broader ascertainment and overidentification of individual deficits. This pattern was particularly evident for functional deficits: identification of falls increased from 8.6% with the standard eFI to 32.8% with LLM augmentation, closer to the 46.7% identified by manual chart review, whereas gait abnormality was identified more frequently by the LLM-enhanced eFI than by manual chart review (54.2% vs 31.7%). These findings are consistent with our prior validation study, in which incorporation of PT-derived information increased ascertainment of functional deficits but also introduced some false-positive classification.^23^

These findings have implications for implementation. LLM augmentation may improve frailty ascertainment more than prognostic discrimination, and its clinical value may therefore depend on the intended use. When the objective is broad screening to identify patients for further geriatric or functional assessment, more complete identification of vulnerability may be valuable despite lower specificity. Conversely, our findings provide limited evidence that LLM augmentation alone improves prediction of adverse post- hospitalization outcomes beyond a primarily diagnosis-coded based eFI. The added complexity of LLM-based extraction should therefore be justified by whether the additional patients identified as frail constitute a clinically actionable population. Future evaluations should examine the clinical utility, resource requirements, and outcomes of acting on enhanced frailty information.^43,48^

This study has several strengths. We evaluated a previously validated LLM- enhanced frailty measure across two distinct hospitalized populations, examining both frailty ascertainment and prognostic performance. Analyses assessed complementary dimensions of performance, including associations with adverse outcomes, discrimination, and threshold-based classification, supported by multiple prespecified sensitivity analyses.

Restricting analyses to hospitalizations with available PT documentation also enabled direct comparison of the standard and LLM-enhanced eFI among patients for whom additional functional information was available, reducing concerns that observed differences reflected selection for PT evaluation alone.

Several limitations warrant consideration. First, frailty-related deficits extracted from the initial PT consultation may reflect baseline vulnerability, acute illness-related functional decline, or both, limiting our ability to distinguish chronic frailty from transient impairment. Furthermore, we used a single-generation LLM (GPT-4o) applied to a single clinical note. More advanced models and approaches incorporating longitudinal or multimodal clinical information, particularly prehospital functional status, may better distinguish chronic vulnerability from acute impairment and yield different prognostic performance. Second, PT documentation was more common among hospitalizations associated with adverse outcomes, raising the possibility of confounding by indication and selection bias. Although findings were generally consistent in analyses restricted to hospitalizations with PT documentation, this approach does not fully address selection bias or establish performance among patients without PT evaluation.

Third, the source cohorts differed in eligibility criteria and clinical context, including the exclusion of index-hospitalization facility discharges from one cohort. Although we adjusted for source cohort and observed broadly consistent findings across cohort-specific and prespecified sensitivity analyses, residual differences may have influenced results. Fourth, approximately one-third of hospitalizations involved middle-aged adults, whereas eFIs have primarily been developed and validated in older populations. However, frailty may capture clinically relevant and unrecognized vulnerability in middle-aged adults with multimorbidity, and findings were similar when restricted to adults aged 65 years or older.

Finally, this single-health-system study may have limited generalizability, as institutional differences in patient populations, PT referral and documentation practices, ICD-10 coding, and availability of clinical narratives could affect the performance of both standard and LLM- enhanced eFIs.

In conclusion, LLM augmentation substantially increased frailty ascertainment but did not significantly improve prognostic discrimination. Improved electronic measurement of frailty and prediction of adverse outcomes should therefore be considered distinct objectives. The value of LLM-enhanced frailty assessment may lie principally in more complete identification of functional vulnerability. Future studies should determine whether the additional patients identified constitute a clinically actionable population and whether using this information to guide care improves outcomes.

## CONTRIBUTORSHIP

All authors made substantial contributions to the conception or design of the work; acquisition, analysis, or interpretation of data; and/or drafting or critical revision of the manuscript. All authors approved the final manuscript and agree to be accountable for the work.

## HUMAN ETHICS & CONSENT TO PARTICIPATE

The study was approved by the Mass General Brigham Institutional Review Board (IRB protocol numbers: **2021P002593** and **2022P001284**). Consent procedures for each source study were approved by the Institutional Review Board.

## USE OF ARTIFICIAL INTELLIGENCe

GPT-4o (OpenAI, deployed through Microsoft Azure) was used as part of the study methodology to extract prespecified frailty-related deficits from physical therapy documentation, as detailed in the Methods and Supplementary Appendix. Generative AI tools were additionally used to assist with manuscript editing and refinement of data visualizations. Figures 1 through 3 were initially generated from study data using R code and subsequently refined with Microsoft Copilot powered by GPT-5.6.

Generative AI did not generate or alter the underlying study data or determine the study results. All AI-assisted outputs were reviewed and verified by the authors, who take responsibility for the final manuscript.

## Data Availability

All data produced in the present work are contained in the manuscript

